# Self-Harm by nurses and midwives: a study of hospital presentations

**DOI:** 10.1101/2022.11.07.22280241

**Authors:** Samantha Groves, Karen Lascelles, Liz Bale, Fiona Brand, Deborah Casey, Keith Hawton

## Abstract

**Background:** Nursing professionals are an occupational group at increased risk of suicide, but little is known about self-harm in this population.

**Aims:** To investigate the characteristics of nurses and midwives who present to hospital following self-harm.

**Method:** We used data from the Oxford Monitoring System for Self-Harm to identify nurses and midwives who presented to the general hospital in Oxford during 2010-2020 following an episode of self-harm and received a psychosocial assessment.

**Results:** During the eleven-year study period, 107 presentations of self-harm involving 81 nurses and midwives were identified. Self-poisoning was the most common self-harm method, with antidepressants and paracetamol most frequently involved. A large proportion had consumed alcohol prior to or during the self-harm act. Some individuals had high suicide intent. Common issues preceding self-harm included problems with a partner, psychiatric disorder, problems with employment, and alcohol misuse. A range of aftercare options were offered following hospital presentation.

**Limitations:** This study was limited to data from a single hospital.

**Conclusion:** Prevention and management of self-harm within this occupational group requires preventative strategies and availability of interventions addressing the range of factors that may contribute to self-harm, especially relationship problems, psychiatric disorders, employment problems, and alcohol misuse.

## INTRODUCTION

Research in several countries, including the United Kingdom (UK) and the United States of America, has shown that nurses, particularly female nurses, are at increased risk of suicide compared to the general population or other occupational groups (e.g., Windsor-Shellard & Gunnell, 2019; Davidson et al. 2020). Concern about suicide among nurses has increased during the COVID-19 pandemic (Rahman & Plummer, 2020). Despite non-fatal self-harm being strongly linked to suicide risk (Carroll, Metcalfe, & Gunnell, 2014), especially in the period shortly after a hospital-presenting self-harm episode (Geulayov et al. 2019), there has been little research on self-harm in nurses. Of a sample of nurses in Hong Kong, 9.3% reported self-harm in the previous year, most commonly by cutting, self-hitting or poisoning (Cheung & Yip, 2016). Factors associated with self-harm were having less than 10 years of clinical experience, chronic illness in the past year, relationship crises with the nurses’ family, and family history of self-harm, together with smoking, psychiatric disorder, and symptoms of stress (Cheung & Yip, 2016). To the authors’ knowledge, self-harm among nurses in the UK (or most other countries) has not yet been examined.

We have utilised a well-established general hospital self-harm monitoring system to identify and study a series of nurses and midwives who presented to hospital following self-harm between 2010 and 2020. The aim was to investigate the characteristics of nurses and midwives who self-harm, including their demographic characteristics, methods of self-harm, problems preceding self-harm and aftercare offered.

## METHODS

### Study sample

The study population included current or former nurses and midwives who presented to the general hospital in [area] between 1^st^ Jan 2010 and the 31^st^ December 2020 following an episode of self-harm and received a psychosocial assessment. Nursing and midwifery students were not included due to inconsistency in reporting of students’ programmes of study within medical records.

### Data Collection

Data were collected using the long-established [area] Monitoring System for Self-harm (e.g., Hawton et al. 2015). Self-harm includes non-fatal intentional self-poisoning, self-injury, or a combination of these methods, irrespective of whether or not there is evidence that that act was intended to result in death (Hawton et al. 2003). Self-poisoning is defined as the intentional self-administration of more than the prescribed dose of any drug. It also includes poisoning with non-ingestible substances and gas. Overdoses of recreational drugs and severe alcohol intoxication are included where the clinical staff consider these are cases of self-harm. Self-injury is defined as any injury that has been deliberately self-inflicted.

Patients who present at the emergency department in the general hospital in [Area] following an episode of self-harm are usually assessed by a specialist mental health clinician and data gathered concerning their sociodemographic (e.g., sex, age, occupation) and clinical characteristics (e.g., psychiatric history, prior self-harm, alcohol misuse), self-reported life problems they are experiencing (e.g., physical health conditions, interpersonal problems), together with a record of what aftercare is offered. Staff usually also complete the Beck Suicide Intent Scale; a 15-item measure examining the severity of suicide intention in relation to self-harm (Beck, Schuyler, & Herman, 1974; Harriss, Hawton, & Zahl, 2005) The scale is comprised of two parts: the first, which involves eight items, assesses the objective ‘circumstances’ of the self-harm act, and the second, is a seven-item ‘self-report’ assessment of the patient’s thoughts and feelings at the time of self-harm. Each item has a score range of 0-2, with a maximum score of 30. For this study, intent was classified as low (score 0-6), moderate (7-12), high (13-20) and very high (21+) (e.g., Hawton, Bale, & Casey, 2021). Drug and alcohol intake are also assessed, with alcohol misuse classified as dependency (with and without physical symptoms) or excessive use (greater than the maximum recommended number of units per week).

Data were extracted for all assessed presentations over an eleven-year period (2010-2020) where the individual’s occupation was coded as a current or former nurse or midwife. These data were merged from two databases, the main monitoring database, which at the time of identifying data for the study included presentations in 2010-2017, and an additional database for which data for 2018-2020 were obtained through hand-searching of monitoring system data forms not yet entered into the main database. After merging of the databases, the data were cleaned.

### Data Analysis

Data were analysed using SPSS version 22 for Windows. Descriptive analyses were conducted. Data are presented as frequencies and percentages. In a minority of cases there were missing data due to incomplete information recorded during psychosocial assessment. Variables with missing data are highlighted within the results. Where individuals presented following multiple self-harm episodes during the study period, the first episode was used as the index.

### Ethical approval

The [Area] Self-Harm Monitoring System has approval to collect data on self-harm for local monitoring and research purposes. The system is compliant with the General Data Protection Regulation (2018) and has approval under Section 251 of the National Health Service (NHS) Act (2006) to collect patient-identifiable information without explicit patient consent.

## RESULTS

### Sample

Across the eleven-year study period (2010-2020), 81 current or former nurses (N= 74) and midwives (N=7) presented to the hospital following 107 episodes of self-harm (Table 1). Most individuals (N=67, 82.7%) had one presentation to the hospital during the study period. Some had repeat presentations (N=14, 17.3%), the numbers of repeats ranging from two to six episodes. Across the study period, numbers of first presentations per year varied (median 8, 3-11 range), including a relatively small number in 2020 (N=5), the first year of the COVID-19 pandemic.

**Table 1.** Characteristics of nurses and midwives who presented with self-harm, by occupation, sex and marital status.

| Characteristic | Employed<br>N= 47 | Unemployed<br>N= 18 | Sick<br>N= 5 | Retired<br>N= 8 | In other<br>employment<br>N= 3 | Total<br>N=81 |
| --- | --- | --- | --- | --- | --- | --- |
| Profession N (%) |  |  |  |  |  |  |
| Nurse | 46 (56.8%) | 15 (18.5%) | 4 (4.9%) | 6 (7.4%) | 3 (3.7%) | 74 (91.4%) |
| Midwife | 1 (1.2%) | 3 (3.7%) | 1 (1.2%) | 2 (2.5%) | 0 (0.0%) | 7 (8.6%) |
| Sex N (%) |  |  |  |  |  |  |
| Female | 44 (54.3%) | 15 (18.5%) | 5 (6.2%) | 8 (9.9%) | 3 (3.7%) | 75 (92.6%) |
| Male | 3 (3.7%) | 3 (3.7%) | 0 (0.0%) | 0 (0.0%) | 0 (0.0%) | 6 (7.4%) |
| Marital Status N (%) |  |  |  |  |  |  |
| Single | 24 (29.6%) | 7 (8.6%) | 3 (3.7%) | 2 (2.5%) | 2 (2.5%) | 38 (46.9%) |
| Married | 13 (16.0%) | 6 (7.4%) | 1 (1.2%) | 4 (4.9%) | 1 (1.2%) | 25 (30.9%) |
| Separated/Divorced | 10 (12.3%) | 5 (6.2%) | 1 (1.2%) | 1 (1.2%) | 0 (0.0%) | 17 (21.0%) |
| Widowed | 0 (0.0%) | 0 (0.0%) | 0 (0.0%) | 1 (1.2%) | 0 (0.0%) | 1 (1.2%) |

Most of the individuals were female (75, 92.6%) (see Table 1) and were largely of white ethnicity (75, 92.6%). Their ages ranged from 21 to 93 years, with a mean of 40.6 (SD 14.6), and median of 39.0. The majority of patients were working as current nurses or midwives (46, 56.8%, nurses; 1, 1.2% a midwife) at the time of hospital presentation. However, a substantial proportion were currently unemployed (15, 18.5%, nurses; 3, 3.7% midwives). Others were on sickness leave (4, 4.9% nurses; 1, 1.2% a midwife), working in another position (3, 3.7% nurse; 0 midwives) or retired (6, 7.4% nurses; 2, 2.5% midwives).

### Self-harm methods

The majority of first self-harm presentations involved self-poisoning (N=58, 71.6%), with just over a fifth involving self-injury (N=18, 22.2%). A further small proportion involved both self-poisoning and self-injury in the same episode (N=5, 6.2%).

The group of drugs most frequently used for self-poisoning (including co-occurrence of self-poisoning and self-injury) were antidepressants (24/63, 38.1%), with fluoxetine, mirtazapine and amitriptyline being the individual drugs most used. Pure paracetamol and paracetamol-containing drugs (23/63, 36.5%) were next most frequent, followed by opiates or other recreational drugs (13, 20.6%), minor tranquillizers and hypnotics (12, 19.0%), other ‘over-the-counter’ or prescribed drugs, (10, 15.9%), and other analgesics (including NSAIDs) (9, 14.3%). Mood stabilisers, major tranquilisers, and non-ingestible poisons (including gas) were each used twice (3.2%). For two patients (3.2%), the substance taken was unknown. Many patients used multiple types of drugs within a single episode (24/63, 38.1%), with one individual using five different substances. When restricting analyses to only nurses and midwives currently employed within the profession, the drugs used for self-poisoning were not different.

Of all episodes involving self-injury (N=23), three (13.0%) included the use of multiple methods. Across all episodes of self-injury (including where multiple methods were used within a single episode) self-cutting of the wrist and/or forearm was the most common method (13/23, 56.5%). Several other methods of self-injury were also used including cuts to other areas of the body (5/23, 21.7%) attempted hanging (3/23, 13.0%), and drowning (2/23, 8.9%), with three other methods each used once.

Of patients with multiple self-harm presentations (N=14), half (N=7, 50.0%) used the same broad method of self-harm (self-poisoning or self-injury) in each self-harm episode, five (35.7%) changed methods across episodes, and two (14.3%) moved from using one method of harm to using both self-poisoning and self-injury simultaneously.

Many patients had consumed alcohol within the six hours before self-harming (35/80, 43.8%). A smaller proportion reported using alcohol as part of the episode (19/75, 25.3%). Only one patient was known to be under the influence of recreational drugs at the time of self-harm.

### Psychiatric history

At first presentation during the study period, just over a quarter of patients were in receipt of outpatient or day patient psychiatric care (21/79, 26.6%). One individual was a psychiatric hospital inpatient. Nearly a quarter had received previous inpatient care (18/76, 23.7%), and a half outpatient care (38/75, 50.7%). Half of the patients were identified as having a psychiatric disorder (36/71, 50.7%), with a smaller proportion a personality disorder (13/59, 22.0%).

#### Previous Self-Harm

Over two-thirds of patients had a known history of previous self-harm (54/78, 69.2%), including nearly half with hospital-presenting self-harm (37/76, 48.7%). Where information on recency of previous self-harm was available (N=47), a quarter of patients had self-harmed within the last month (12/47, 25.5%).

#### Substance Misuse

Excessive consumption of alcohol was relatively common among the nurses (18/79, 22.8%), with a few additional cases of alcohol dependency (8/79, 10.1%). Drug use was rare, with only five patients reporting use (6.2%). Across all cases, the most common substances used were opiates (3, 3.7%) and cocaine (2, 2.5%). One patient reported misuse of benzodiazepines and another of cannabis.

### Suicide intent

Data regarding scores on the Suicide Intent Scale were available for 86.4% (N=70) of first presentations. The median suicide intent score was 8.5 (range 0-28). The scores of a third of patients (23/70, 32.9%) indicated low suicide intent (score 0-6), another third (25/70, 35.7%) moderate intent (7-12), one fifth (14/70, 20.0%) high intent (8-20), and eight presentations (11.4%) were classified as involving very high suicide intent (21+).

### Problems faced by individuals

The most common problems nurses and midwives were facing at the time of self-harm were relationship difficulties, these being identified in nearly half the patients (38, 46.9%). The next most common problem was related to psychiatric disorder (24, 29.6%). Approximately a quarter of the patients had problems related to employment (22, 27.2%) and a similar proportion had problems in their relationship with their families (20, 24.7%). No nurses or midwives were identified as having problems concerning sexual adjustment, or as a result of being a carer (see Table 2). At the time of presentation, just over one fifth of patients reported having experienced violence from others in the previous five years (15/72, 20.8%), with a small proportion saying they had been violent towards others (6/74, 8.1%).

**Table 2:** Problems identified in nurses and midwives at assessment after first self-harm presentation.

| Problem | N (%)<sup>a</sup><br>N=81 |
| --- | --- |
| Relationship with partner | 38 (46.9%) |
| Psychiatric disorder | 24 (29.6%) |
| Employment | 22 (27.2%) |
| Relationship with family | 20 (24.7%) |
| Alcohol | 19 (23.5%) |
| Physical health | 16 (19.8%) |
| Finance | 15 (18.5%) |
| Social isolation | 13 (16.0%) |
| Bereavement | 11 (13.6%) |
| Housing | 8 (9.9%) |
| Childhood sexual abuse | 7 (8.6%) |
| Drugs | 7 (8.6%) |
| Eating disorder | 7 (8.6%) |
| Relationship with friends | 6 (7.4%) |
| Childhood emotional abuse or neglect | 4 (4.9%) |
| Childhood physical abuse | 4 (4.9%) |
| Chronic pain | 4 (4.9%) |
| Legal | 2 (2.5%) |
| Repetitive self-mutilation | 2 (2.5%) |
| Bullying<sup>b</sup> | 1 (1.2%) |
| Sexual adjustment | 0 (0.0%) |
| Caring duties<sup>b</sup> | 0 (0.0%) |

<sup>a</sup> Multiple problems were recorded for some patients; therefore percentages exceed 100%.
<sup>b</sup> 19.8% of data missing; measures were added from 2012 onwards.

A large proportion of patients reported physical health conditions (33/78, 42.3%), with the most common conditions including musculoskeletal (7/78, 9.0%), respiratory (6/78, 7.7%), and gynaecological problems (5/78, 6.4%). Multiple health conditions were reported by ten patients (10/78, 12.8%).

### Aftercare Offered

Following their first presentation, approximately half of the patients were offered outpatient care (41, 50.6%) and nine (11.1%) were admitted to psychiatric inpatient care. A small proportion were referred to the ‘*Improving Access to Psychological Therapies’* (IAPT) national psychological counselling programme (8, 9.9%), at times alongside other aftercare. Over a third of patients were referred back to their general practitioners, (30, 37.0%); for some patients this was alongside other aftercare offers (12 /30, 40.0%). Small proportions of patients were referred to alcohol services (6, 7.4%), or social services (2, 2.5%).

## DISCUSSION

We examined characteristics of nurses and midwives who presented to hospital following self-harm over an eleven-year period. The vast majority of individuals were female (92.6%). While we were unable to estimate the relative rates of self-harm by sex, it is well recognised that risk of suicide is particularly elevated in female nurses in the UK (Windsor-Shellard & Gunnell, 2019). The final year of the study period saw the beginning of the COVID-19 pandemic. This may have been a reason for the relatively small number of self-harm presentations in 2020 as hospital presentations for self-harm in general were reduced during the initial phase of the pandemic (e.g., Hawton et al. 2021).

The majority of self-harm episodes involved self-poisoning. This is consistent with research on suicide (e.g., Hawton et al. 2011) and suicide attempts (Braquehais et al. 2016) among nurses. However, greater use of poisoning was only found in female nurses in one study (Davidson et al. 2020). The common use of self-poisoning contrasts with a self-report study of self-harm among nurses, where cutting was most frequent, followed by self-hitting and then poisoning (Cheung & Yip, 2016). This difference may be a result of the previous study including self-reported self-harm, not just hospital presentations. Self-cutting is far more common in self-harm episodes in the community, whereas self-poisoning is more frequent in individuals presenting to hospitals, at least in young people (Geulayov et al. 2018).

As in the current study, antidepressants, paracetamol (acetaminophen), and opiates or other recreational drugs have been shown to be commonly used for self-poisoning among nurses (Davidson et al. 2020; Choflet et al. 2021). In the present sample, the frequent use of antidepressants for self-harm may reflect the fact that many of the individuals were experiencing psychiatric disorders, especially depression, at the time of self-harm, as found in nurses who died by suicide (Hawton et al. 2002). The use of opiates (recreational and non-recreational) may be linked to prevalence of musculoskeletal or pain-related conditions, for which nurses are at a high risk (Davis & Kotowski, 2018).

The present study contributes to the discussion of whether self-poisoning among nursing professionals is related to access to medication in the workplace, and/or knowledge regarding lethality. Like other studies examining suicide in nurses (Davidson et al. 2020), prescribed or over-the-counter medication (other than recreational opiates) were most frequently used, suggesting workplace access is not often a contributor, unlike suicidal behaviour in other health-related professions such as anaesthetists (e.g., Plunkett et al. 2021).

Alcohol was frequently used prior to and as part of the self-harm acts, and alcohol misuse was relatively common among the nurses. Possible reasons for excessive alcohol use may include work stress (Foli et al. 2020) and use as a sleep aid (Dorrian et al. 2011). In the assessment of nursing staff who have self-harmed or are thought to be at risk, problematic alcohol use should be considered given research demonstrating high levels of problematic alcohol use among nurses who have died by suicide (e.g., Hawton et al. 2002), and use of alcohol as part of suicide acts (e.g., Choflet et al. 2021).

The proportion of the nurses and midwives with a history of previous self-harm was considerable, with over two-thirds having at least one previous episode. Previous episodes were often relatively recent (most not involving hospital presentation). Furthermore, repeat episodes leading to further hospital presentation during the study period were quite common. This is particularly concerning given the association between prior self-harm and later suicide among nurses and midwives (Hawton et al. 2002), the known increased risk of suicide associated with repetition of self-harm (Geulayov et al. 2019), and the substantial proportion of self-harm presentations in the present study with high or very high suicidal intent.

The majority of the nurses and midwives had previous contact with psychiatric services, and a large proportion had psychiatric disorders. While we didn’t have detailed information about specific conditions, anxiety, depression, and panic disorder have been linked to suicidal behaviours among nurses (e.g., Stelnicki et al. 2020). This highlights the need for appropriate support and treatment for nurses experiencing mental health conditions.

Relationship difficulties were the most common problem identified by nurses and midwives. This correlates with other studies of self-harm (Cheung & Yip, 2016) and suicide among nurses (e.g., Hawton et al. 2002). Employment-related issues were also highly prevalent among nurses and midwives. Specific problems may include job loss, work-related stress, and stigma related to seeking support regarding suicidality (Wolf et al. 2020; Davidson et al. 2021). Physical health problems and comorbidity were common within the sample.

Reasons for this, particularly regarding musculoskeletal conditions, may include high physical demands of nursing, for example, frequent lifting and long periods spent standing. Health problems including chronic pain have been found to contribute to suicide in nurses (e.g., Davidson et al. 2021).

### Strengths and Limitations

The study findings are based on detailed and systematic clinical assessments, with the method of data collection being consistent across the study period. However, detailed information was only available on nurses who had a psychosocial assessment. Nurses and midwives who self-harmed in the community and did not present to hospital will not have been included in the study. Their characteristics, including method used for self-harm, are likely to differ from those of nurses and midwives presenting to hospital following self-harm (Cheung & Yip, 2016).

The information attained for this study was largely based on a single clinical assessment. Also, we only had information on aftercare offered, not received. Finally, the results of the study are limited by being from a single centre, in a relatively affluent area (Geulayov et al. 2019). Participants of the study were largely of white ethnicity (92.6%), a higher proportion than [area’s] population (78%) ([area] City Council, 2011) and the NHS non-medical workforce, in which nursing is included (80.3%) (NHS Workforce, 2021).

### Implications for Research and Policy

Further research should explore the nature of employment problems that nurses may identify as precedents to self-harm, for which qualitative approaches may be most appropriate. In addition, although case-control studies have been conducted examining characteristics of nurses who have died by suicide (e.g., Hawton et al. 2002), to the authors’ knowledge, characteristics of those self-harming have not been examined in this manner. The use of a case-control study comparing characteristics of self-harm among nursing professionals with the general population or a reference occupation could clarify whether any factors differ, and thus may be particularly important for targeted interventions for self-harm and suicide prevention for nurses. A longitudinal study in which nurses and midwives who self-harm are followed up to assess aftercare received and its outcome would provide further understanding of the significance of self-harm in this population.

The findings of the present study have highlighted factors that may inform primary and secondary prevention initiatives to target problems nurses and midwives might face, in turn contributing to prevention of self-harm and suicide. Primary initiatives should include education regarding stress management, injury prevention, and safe levels of alcohol use. Secondary intervention initiatives could include support for nurses and midwives with harmful levels of alcohol use, ensuring continuous learning opportunities regarding moving and handling, tailored health promotion resources to increase awareness and reduce stigma surrounding mental health conditions, flexible and accessible workplace counselling and robust clinical supervision arrangements.

## Data Availability

Research data are not shared.

